# Cross-sectional and longitudinal genotype to phenotype surveillance of SARS-CoV-2 variants over the first four years of the COVID-19 pandemic

**DOI:** 10.1101/2024.04.18.24305862

**Authors:** A Akerman, C Fichter, V Milogiannakis, C Esneau, MR Silva, T Ison, JA Lopez, Z Naing, J Caguicla, S Amatayakul-Chantler, N Roth, S Manni, T Hauser, T Barnes, T Boss, A Condylios, M Yeang, K Sato, NW Bartlett, D Darley, G Matthews, DJ Stark, S Promsri, WD Rawlinson, B Murrell, AD Kelleher, DE Dwyer, V Sintchenko, J Kok, S Ellis, K Marris, E Knight, VC Hoad, DO Irving, I Gosbell, F Brilot, J Wood, A Aggarwal, SG Turville

## Abstract

**Background:** Continued phenotyping and ongoing surveillance are important in current and future monitoring of emerging SARS-CoV-2 lineages. Herein we developed pragmatic strategies to track the emergence, spread and phenotype of SARS-CoV-2 variants in Australia in an era of decreasing diagnostic PCR testing and focused cohort-based studies. This was aligned to longitudinal studies that span 4 years of the COVID-19 pandemic.

**Methods:** Throughout 2023, we partnered with diagnostic pathology providers and pathogen genomics teams to identify relevant emerging or circulating variants in the New South Wales (NSW) community. We monitored emerging variants through viral culture, growth algorithms, neutralization responses and change entry requirements defined by ACE2 and TMPRSS2 receptor use. To frame this in the context of the pandemic stage, we continued to longitudinally track neutralisation responses at the population level using using sequential batches of pooled Intravenous Immunoglobulins (IVIG) derived from in excess of 700,000 donations.

**Findings:** In antibodies derived from recent individual donations and thousands of donations pooled in IVIGs, we observed continued neutralization across prior and emerging variants with EG.5.1, HV.1, XCT and JN.1 ranked as the most evasive SARS-CoV-2 variants. Changes in the type I antibody site at Spike positions 452, 455 and 456 were associated with lowered neutralization responses in XBB lineages. In longitudinal tracking of population immunity spanning three years, we observed continued maturation of neutralization breadth to all SARS-CoV-2 variants over time. Whilst neutralization responses initially displayed high levels of imprinting towards Ancestral and early pre-Omicron lineages, this was slowly countered by increased cross reactive breadth to all variants. We predicted JN.1 to have a significant transmission advantage in late 2023 and this eventuated globally at the start of 2024. We could not attributed this advantage to neutralization resistance but rather propose that this growth advantage arises from the preferential utilization of TMPRSS2 cleavage-resistant ACE2.

**Interpretation:** The emergence of many SARS-CoV-2 lineages documented at the end of 2023 to be initially associated with lowered neutralization responses. This continued to be countered by the gradual maturation of cross reactive neutralization responses over time. The later appearance and dominance of the divergent JN.1 lineage cannot be attributed to a lack of neutralization responses alone, and we support its dominance to be the culmination of both lowered neutralization and changes in ACE2/TMPRSS2 entry preferences.

## Introduction

We have now entered a phase of the coronavirus disease 2019 (COVID-19) pandemic in which community immunity is being maintained through a combination of decreasing vaccine doses coupled with ongoing infection waves (https://ourworldindata.org/coronavirus). Whilst global efforts to monitor daily COVID-19 case numbers and vaccine doses have been significantly reduced, there remains a need for sustainable surveillance of emerging SARS-CoV-2 variants at the genomic and phenotypic levels. This continued effort, as with other viral epidemic surveillance, allows rapid identification of any shifts within the virus’s ability to evade immune responses and/or entry requirements [3] to enable public health responses, if required.

At the start of the pandemic, access to circulating variants could be readily achieved through diagnostic-research partnerships [3] by providing access to remnant diagnostic PCR-positive samples for viral isolation and whole genome sequencing. Following the introduction of Rapid Antigen Tests (RATs) and reduction of diagnostic PCR testing, the access to remnant swabs for rapid phenotypic surveillance was significantly reduced. In response to these changes, we established a genotype to phenotype surveillance networkIn addition to routine genomic surveillance conducted in NSW, national and global monitoring of variant prevalence and growth competition was inferred using the global GISAID datasets (https://github.com/MurrellGroup/lineages) and this then enabled a pragmatic means to short-list key emerging variants for testing. Phenotypic surveillance was then linked to genomic surveillance with parallel isolation and phenotyping platforms that enabled rapid resolution of viral phenotypes in terms of neutralization resistance, and rapid analysis of viral entry requirements at the cell surface for current variants. We then linked these observations to longitudinal studies monitoring neutralization responses and entry factors for variants that have dominated the first three years of the pandemic; 2020 through to late 2023. The inclusion of the latter longitudinal neutralisation datasets was important at several levels. Firstly, it established a trajectory following key immunological events (vaccination doses and/or infection caseloads). Secondly, it provided an immunological history that frames current responses. Finally it also addressed concomitent evolving viral entry requirements over this same time period.

Herein whilst we observe continued resistance to neutralization responses in contemporary variants such as HK.3, XCT and JN.1, longitudinal datasets observe maturation of the host response over time to counter these viral gains. Furthermore, we observed continued evolution of the virus to favour entry requirements that could be readily and rapidly observed using clonal cell lines with and without TMPRSS2 cleavage sensitive ACE2. Overall these datasets observe two major trajectories over the first 3 to 4 years of the pandemic. Firstly, whilst maturation of neutralization responses has been at a slower rate than the emergency of variant resistance, it is creating continued pressure on emerging variants. Secondly, whilst the entry phenotypes of early SARS-CoV-2 mirrored that of SARS-CoV-1 and are defined by increased replication in the presence of TMPRSS2-ACE2 cleavage, continued evolution of SARS-CoV-2 has progressively consolidated away from this pathway and now primarily uses ACE2 that is resistant to TMPRSS2 cleavage.

## Methods

### Polyclonal immunoglobulin preparations and anti-SARS-CoV-2 hyperimmune globulin

Polyclonal IgG batches were purified using the licensed and fully validated immunoglobulin manufacturing process used for Privigen [4]. Privigen batches were manufactured using the Privigen process [4] and included U.S. plasma collected by plasmapheresis from a mixture of vaccinated (SARS-CoV-2 mRNA vaccines), convalescent and non-convalescent donors (source plasma, approximately 15,000 donations per batch) collected over the period of May 2020 to June 2023.

### Human plasma

Cohort samples with linked vaccine status and infection history information (Supplementary Table S1), collected within the first half of 2023, were used to complement data from IVIG testing. The concluding ADAPT cohort [5] consisted of recent donations in 2023 from a group that were vaccinated and boosted with a documented with a BA.1 infection. The latter is consistent with a significant proportion of the community within the targeted surveillance state of NSW.The Australian Red Cross Lifeblood (Lifeblood) cohort (LIFE) comprised of 44 plasma samples collected from volunteers presenting to Lifeblood for whole blood or plasma donations. The donors included in this study were both infected and vaccinated for SARS-CoV-2 with individuals having received a minimum of two vaccine doses and reporting an infection between August 2022 and March 2023. Infections were confirmed by a Rapid Antigen Test.

### Cell culture

HEK293T cells stably expressing human ACE2 and TMPRSS2 were generated by lentiviral transductions as previously described [23, 24]. A highly permissive clone (HAT-24) was identified through clonal selection and used for this study. The HAT-24 cell line has been extensively cross-validated with the VeroE6 cell line [23]. VeroE6-TMPRSS2 (Vero-T) cells were kindly provided by Professor Alex Khromykh (University of Queensland). HAT-24 and Vero-T cells were cultured in Dulbecco’s Modified Eagle Medium (Gibco, 11995073) containing 10% foetal bovine serum (Gibco, 10099141; DMEM-10%FBS). All cells were incubated at 37°C, 5% CO2 and >90% relative humidity. For the Vero-T cell line, authentication was performed as previously described [23, 24]. The STR profiling of HAT-24 has been previously described [23]. All cell lines tested negative for mycoplasma. Primary bronchial epithelial cells (pBEC) were provided by P. A. B. Wark (University of Newcastle), and originally obtained during bronchoscopy, with written informed consent. Experiments were conducted with approval from the University of Newcastle Safety Committee (Safety REF# 25/2016 and R5/2017). All participants underwent fibre-optic bronchoscopy in accordance with standard guideline [6]. pBEC cultures were grown and differentiated until confluent in complete Bronchial Epithelial Cell Growth Basal Medium (Lonza, CC-3171) before use for air–liquid interface (ALI) experiments. All cells were cultured and incubated at 37 °C, 5% CO2 and >90% relative humidity, unless otherwise indicated.

### Virus isolation, expansion, and titration

All laboratory work involving infectious SARS-CoV-2 was undertaken in biosafety level 3 (BSL-3) conditions under existing approved safety protocols. SARS-CoV-2 variants were isolated and characterized as previously described [18, 19]. In brief, primary diagnostic nasopharyngeal swabs that are RT-qPCR-positive for SARS-CoV-2 were sterile filtered through a 0.22 µm column filter at 10,000 *xg*, serially diluted using a 1:3 series then transferred to HAT-24 cells (5 x 10^3^/well in 384-well plates). Supernatant (300 µL; passage 1) from infected wells (positive for cytopathic effects via light microscopy) was transferred to 0.5 x 10^6^ Vero-T cells in a 6-well plate (in 2 mL of MEM-2%FBS) and incubated under standard culture conditions (37°C, 5% CO_2_) until significant cytopathic effects were observed (24-72 hours). Supernatant (passage 2) was collected and cleared via centrifugation at 3000 rpm for 10 minutes, frozen at –80°C then thawed to determine median 50% tissue culture infectious dose (TCID_50_/mL) in Vero-T cells, according to the Spearman-Karber method [25]. To generate passage 3 viral stocks, Vero-T cells were infected at MOI = 0.025, incubated for 24 hours and the supernatant collected, cleared and frozen as for passage 2 stocks. Sequence identity and integrity were confirmed for both passage 1 and passage 3 virus via whole-genome viral sequencing using Oxford Nanopore technology platform, as previously described [19, 26]. For a list of the viral variants isolated in this study, see Supplementary Table 1. Passage 3 stocks were titrated by serial dilution (1:5 series) in DMEM-5%FBS, transferred to HAT-24 cells live-stained with 5% v/v nuclear dye (Invitrogen, R37605) at 1.6 × 10^4^ cells/well in 384-well plates and incubated for 20 hours. Whole-well nuclei counts were determined using an IN Cell Analyzer 2500HS high-content microscope and IN Carta analysis software (Cytiva, USA). Data was normalized to generate sigmoidal dose–response curves (average counts for mock-infected controls = 100%, and average counts for highest viral concentration = 0%) and median Virus Effective (VE_50_) values were obtained with GraphPad Prism software.

### Abbott Architect SARS-CoV-2 anti-Nucleocapsid IgG

IgG antibodies to the nucleocapsid protein (N) of SARS-CoV-2 were detected using Architect SARS-CoV-2 IgG (Abbott Diagnostics, Sydney, NSW Australia) as previously described [21].

### Flow cytometry cell-based assay for detection of SARS-CoV-2 antibodies

A flow cytometry cell-based assay detected patient serum antibodies against SARS-CoV-2 antigens as previously described [5]. SARS-CoV-2 full-length Spike (Wuhan-1) were transiently expressed on transfected HEK293 cells. Serum (1:80) was added to live Spike-expressing cells followed by AlexaFluor 647-conjugated anti-human IgG (H+L) (Thermo Fisher Scientific). Cells were acquired on the LSRII flow cytometer (BD Biosciences, USA). The assay was verified using 10 antibody standards from the National Institute for Biological Standards and Control (NIBSC) distributed during the CS678 protocol for the World Health Organization (WHO) collaborative study to establish the first International Standard for anti-SARS-CoV-2 antibody and Reference Panel. Data were analyzed using FlowJo 10.4.1 (TreeStar, USA), Excel (Microsoft, USA), and GraphPad Prism (GraphPad Software, USA).

### R-20: Rapid, high-content SARS-CoV-2 microneutralization assay with HAT-24 cells

The R-20 high-content neutralization assay was performed using HAT-24 cells, as previously described [18, 19, 23]. In brief, human plasma or IVIGs were serially diluted using a 1:2 series in DMEM-5% starting at 1:10 (dilution of samples tested against ancestral Clade A.2.2 started as 1:40). SARS-CoV-2 virus (diluted in the respective media) standardized to 2x VE_50_ was added to diluted samples and this virus-serum/antibody mixture was incubated at 37°C, 5% CO_2_ for 1 hour. 40 µL (in technical duplicates) was transferred to pre-plated nuclear stained HAT-24 cells (1.6 x 10^4^/well) in 384-well plates. Plates were incubated for 20 hours, and cell nuclei enumerated using an IN Cell Analyzer 2500HS high-content microscope. The % neutralization was calculated with the formula: %N = (D − (1 − Q)) × 100/D as previously described [24]. “Q” is a well’s nuclei count divided by the average count for uninfected controls (defined as having 100% neutralization) and D = 1 − Q for the average count of positive infection controls (defined as having 0% neutralization). Sigmoidal dose–response curves and IC_50_ values (reciprocal dilution at which 50% neutralization is achieved) were obtained with GraphPad Prism software.

### Cleavage sensitive and cleavage resistant entry assays using primary SARS-CoV-2

TMPRSS2 cleavage sensitive (WT ACE2 +TMPRSS2) and cleavage resistant ACE2 (NC-ACE2 + TMPRSS2) were generated as previously described [7]. Virus titrations were carried out by serially diluting expanded viral stocks (1:5) in MEM-2%FBS, mixing with cells initially in suspension at 5 × 10^3^ cells/well in 384-well plates and then further incubating for 72 hours. The cells were then stained live with 5% v/v nuclear dye (Invitrogen, R37605) and whole-well nuclei counts were determined with an IN Cell Analyzer 2500HS high-content microscope and IN Carta analysis software (Cytiva, USA). Data was normalized to generate sigmoidal dose– response curves (average counts for mock-infected controls = 100%, and average counts for highest viral concentration = 0%) and median Virus Effective (VE_50_) values were obtained with GraphPad Prism software

### Statistics

Statistical analyses were performed using GraphPad Prism 9 (GraphPad software, USA). Sigmoidal dose response curves and interpolated IC_50_ values were determined using Sigmoidal, 4PL model of regression analysis in GraphPad Prism. For statistical significance, the datasets were initially assessed for Gaussian distribution, based on which further analysis was performed. For datasets that followed normal distribution One way ANOVA was used while for others non parametric tests such as Friedman (for paired samples) or Kruskal Wallis test (for unpaired samples) with Dunn’s multiple comparison were employed. In all cases the data was compared to the Ancestral Clade A.2.2. Mann Whitney U test was used to analyze data between two groups. Details of statistical tests used for different data sets have also been provided in figure legends.

### Ethics

Human research ethics approval for this study was granted by Lifeblood Research Ethics Committee (30042020) and for the ADAPT cohort St Vincent’s Hospital (2020/<u>ETH00964</u>) with donor consent forms including a statement that blood donations may be used for research purposes. NSW Health facilitated provision of de-identified residual COVID-19 diagnostic swabs for use in this study to support public health response under the governance of Health Protection NSW.

### Sample Collection

Widespread PCR COVID-19 testing gave unparalleled access to samples for viral isolation throughout the pandemic. With the emergence of Rapid Antigen Tests (RATs) and reduced access to PCR testing for the general population, sample access for phenotypic analysis was reduced. To enable the continuation of timely phenotypic surveillance of emerging variants, a collaboration between the NSW Ministry of Health (MoH), NSW pathogen genomics reference laboratory, diagnostic pathology providers and the Kirby Institute was established. Swab samples were collected from either a single diagnostic centre in Sydney that tests samples accrued from 20 COVID-19 collection sites covering the Sydney Metropolitan area or the diagnostic laboratory Histopath.

Key to establishing this joint diagnostic-research protocol was initial triaging of samples where diagnostic Ct values were less than 25, aliquoting of a small sample (100 µl) of the remnant swab into a coded tube (to enable patient de-identification), freezing this sample at –80°C within 24 hours and then processing of the remaining sample for WGS. In the first half of 2023 this enabled isolation rates of >70%, with increasing isolation rates with each emerging SARS-CoV-2 variant lineage.

## Results

### Phenotypic surveillance in Australia in early 2023

In first half of 2023, CH.1.1, XBB.1.16, XBB.1.5 and then XBB.1.9-derived lineages dominated the variant mix in Australia (Fig. 1A). Ranking of growth advantages with global WGS datasets revealed the XBB.1.9 derivative EG.5.1 to have a competitive advantage (Fig. 1B). Implementation of the consolidated phenotypic surveillance observed primary virus isolation rates increase over time in early 2023: CH.1.1 lineages = 68% (*n = 50*), XBB.1.5 lineages = 72% (*n = 51*), XBB.1.16 lineages = 74% (*n = 36*), and XBB.1.9 lineages = 82% (*n = 88*). During the isolation phase of phenotyping, infectivity per viral load was calculated in each nasopharyngeal swabs sample to determine if infectivity per Ct value correlated with the appearance of emerging variants. Infectivity of swabs per Ct value was consistent with the growth advantage estimates, with initial XBB.1.9 trending towards increased infectivity per diagnostic Ct value but not significantly different to other early co-circulating lineages (Fig. 1C-D). The progressive acquisition of Spike polymorphisms F456L and L455F (Fig. 1E) at the type I antibody binding site, within XBB.1.9-derived lineages EG.5.1 and HK.3, was then associated with significant increases in infectivity per diagnostic Ct value. This further supported their trajectory to dominance among variants towards the latter half of 2023. Changes at the type I antibody binding site at positions 455 and 456 appear to be related to two phenotypic outcomes, as this is also the Spike-ACE2 binding site [8]. F456L has been shown to lower ACE2 binding, whereas L455F facilitated an increase in ACE2 binding to compensate for the loss through F456L [8]. Whilst this may explain the increased infectivity per Ct value of lineages such as HK.3 (bearing F456L and L455F Spike polymorphisms), the increase of infectivity for EG.5.1, which encompasses only the F456L mutation, could not initially be associated with increases in ACE2 affinity.

**Figure 1.**
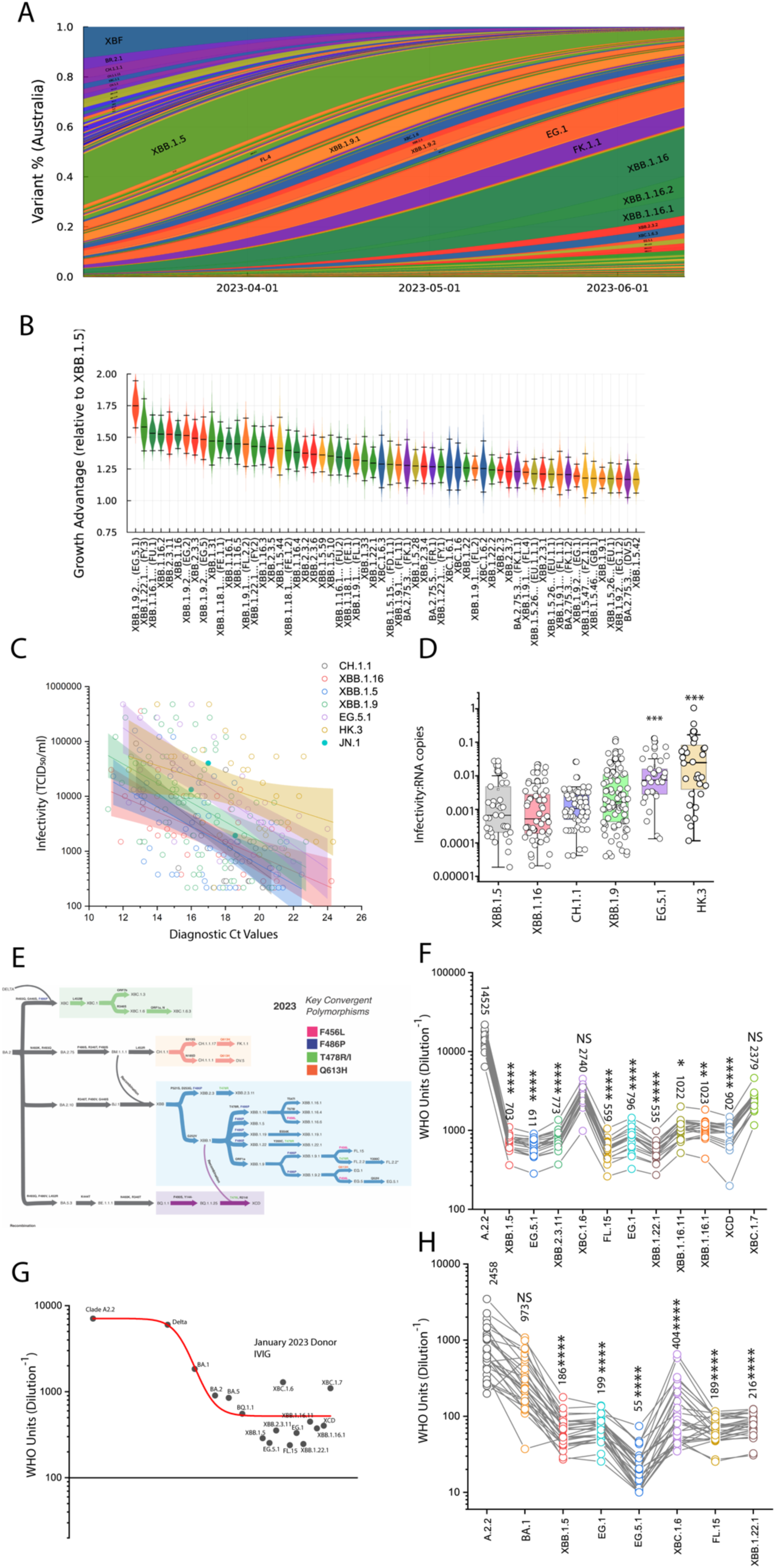
Consolidated model of SARS-CoV-2 surveillance in NSW, Australia and the appearance of convergent XBB lineages in mid 2023. **A.** Surveillance summary of Australia in June 2023 based on genomic data via GISAID. Each vertical slice depicts the posterior mean variant frequency estimate from a hierarchical Bayesian multinomial logistic model of variant competition. Two dominant variant lineages are derived from XBB.1.16 (dark green) and XBB.1.9 (orange). **B.** Results from a model of global SARS-CoV-2 lineage competition. Estimates of multiplicative growth advantage (per week) for lineages are provided, both relative to the parental BA.2, and to the recently dominant XBB.1.5. **C-D.** Summary of over 500 endpoint titers from primary nasopharyngeal swabs covering the diagnostic period of 2023. Lineages are grouped based on nearest parent (CH.1.1, XBB.1.5, XBB.1.9, XBB.1.16) or sub-lineages that have acquired key Spike polymorphisms and sustained high prevalence (XBB.1.9-derived EG.5.1 and HK.3). Samples of JN.1 that appeared during this period (n = 3) are presented as an initial comparison. **C.** Summary plotted as a linear regression of infectivity (TCID50/ml; Y-axis) versus diagnostic Ct value. **D.** RNA copies calculated per diagnostic Ct value were then used to calculate infectivity to RNA ratios to enable statistical comparisons across groups. **E.** From left to right, Omicron lineages from the parent BA.2 are now primarily split across recombinant groupings in Australia by mid-June of 2023. From top to bottom: The top grouping represents the Delta-BA.2 recombinant lineages XBC followed by the non-recombinant CH.1.1-derived lineages. The third group from the top is primarily XBB recombinant lineages covering XBB.2.3, XBB.1.5, XBB.1.9, XBB.1.19 XBB.1.22.1 and the recombinant recombinant XCD. Convergent Spike polymorphisms across lineages are presented in blue (F486P), pink (F456L), green (T478R/I) and orange (Q613H). The bottom group represents BQ.1.1-derived lineages. **F.** Representative panel of emerging variants appearing from early to mid 2023 tested for neutralization responses against 20 IVIG batches manufactured from U.S. donations collected in late 2022 (Supplementary Table SI). **G.** Trajectory of neutralization responses are summarized with the use of the most recent IVIG batches manufactured from donations in 2023. Major variants are presented chronologically until BQ.1.1. Additional emerging variants are presented independent of time of appearance in the community. Lowest neutralization responses are consistent with highest rank variants in B. EG.5.1 and XBB.1.22.1. **H.** Representative panel of emerging variants appearing from early to mid 2023 tested for neutralization responses against 2023 individuals from the ADAPT cohort (Supplementary Table SI). * = p<0.05; ** = p<0.01; *** = p<0.001; **** = p<0.0001.

Given our access to hundreds of primary isolates (Supplentary Tables I), we used a system of triage to focus the testing on key emerging variants. This prioritization used a combination of growth rate calculations (Fig. 1B) and concurrent testing using pooled antibodies to rank samples for testing against large sample cohorts. Twenty batches of clinical grade IVIG was used in this setting as each batch represented 10% IgG (w/v) derived from pools of approximately 15,000 donors per batch over a known donation period covering a range of approximately 4 weeks [9]. We observed lineages such as EG.5.1, FL.15 and XBB.1.22.1 were the most resistant to neutralisation using IVIG batches (Fig. 1F) and this was consistent with EG.5.1 and XBB.1.22.1 lineages leading forcast epidemiological growth advantages in mid-2023 (Fig. 1B). Given IVIG batches have been observed to have increasing neutralising breadth over time [9], we focused our analysis on prior dominant lineages in the pandemic versus those appearing in the mid-half of 2023 (Fig. 1G). Herein, stepwise loss of IVIG titers were observed with the appearance of early Omicrons from BA.2, and XBB.1.5 to the current EG.5.1 and XBB.1.22.1. Consistently through 2022 and 2023, recombinant lineages derived from the Delta-BA.2 recombinant XBC appeared in Australia, which led to neutralization profiles like Omicron BA.2 (Fig. 1G). Using a representative set of variants that covered the continuum of variants tested against IVIG panels, we confirmed similar neutralization profiles at the individual donor level in the ADAPT cohort [5], with the only exception being EG.5.1, which provided the lowest neutralization responses across this group (Fig. 1H).

### Surveillance of longitudinal SARS-CoV-2 population antibody responses from 2021 to 2023 establishes the major immunological events of the pandemic to date

Anti-Spike IgG, Anti-Nucleocapsid IgG, neutralization titers and neutralization breadth over time were analyzed and associated with publically available U.S. datasets on case number surrogates (wastewater viral load surveillance, copies per mL courtesy of Biobot Analytics) and vaccine doses admininistered over time. Monitoring the sum of 688,199 plasma donations collected from September 2021 to January 2023, a time frame that started prior to the peak third dose vaccination roll out in the U.S., we followed events from the resolution of the Delta wave in late 2021 and continued analysis through till January 2023 (Fig. 2A-B). This time frame covered most of the key immunological events such as the combined booster (third) vaccine roll out and Omicron BA.1 wave (the largest wave of infection in the U.S), the Omicron BA.2 wave which coincided with a smaller third vaccine dose rollout, the Omicron BA.5 wave, the approval and release of BA.4/5 bivalent vaccines and the Omicron BQ.1 wave. Here we observed increased levels of anti-Nucleocapsid IgG shortly following the Delta, BA.1, BA.5 and BQ.1 waves (Fig. 2A). Whilst the wastewater viral loads were lower in magnitude, this data demonstrated a similar pattern across the three distinct Omicron infection waves. Interestingly, higher levels of anti-Nucleocapsid IgG were observed across the latter Omicron BA.5 and BQ.1 infection waves, which may reflect immunological recall of a significant proportion of the population being reinfected during those latter waves (Fig. 2A). Live Virus Neutralization Titers (LVNT) against ancestral Clade A.2.2 peaked during the COVID-19 pandemic following the large vaccine booster roll out (Fig. 2B), which coincided with the largest reported U.S. infection wave of Omicron BA.1 (Fig. 2B). Additional lower level LVNT peaks were observed during subsequent booster roll outs, as shown by smaller peaks in LVNT during the third/fourth dose roll out and the introduction of bivalent vaccines (Fig. 2B). To further understand the relationship between LVNT, COVID-19 case numbers, and COVID-19 vaccine doses, we plotted LVNT versus anti-Nucleocapsid IgG (Fig. 2C, E) and LVNT versus anti-Spike IgG (Fig. 2D, F). Through this comparison, we observed no correlation of LVNT and anti-Nucleocapsid IgG (*r = 0.07796; p = 0.55039*). In contrast, LVNT versus anti-Spike IgG (Fig. 2F) were positively correlated (*r = 0.4258*; *p <0.00001*). Overall, the initial vaccine booster dose roll out combined with the large BA.1 infection wave contributed to a level of population immunity revealing peak immune responses of the pandemic began at the start of 2022. Following resolution of this wave, smaller peaks in LVNT were associated with subsequent booster roll outs rather than case waves, resulting in LVNT remaining stable at around 10,000 WHO units/mL to the ancestral Clade A.2.2.

**Figure 2.**
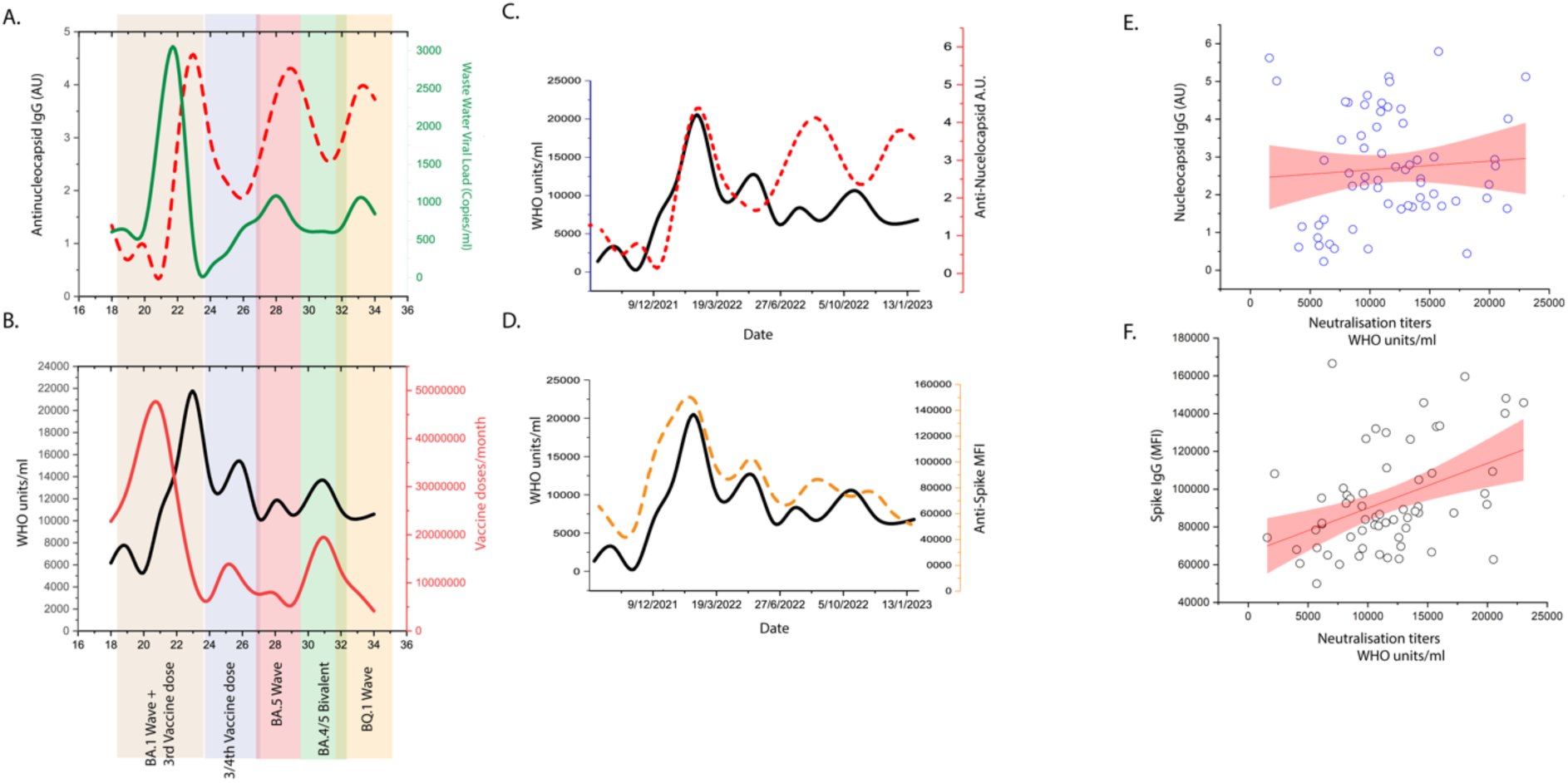
Tracking of neutralization titers across the major immunological events in the U.S. population using pooled intravenous immunoglobulins (IVIGs). **A**. Anti-Nucleocapsid IgG levels (dashed line) in pooled intravenous immunoglobulins (IVIGs) collected throughout the COVID-19 pandemic, commencing at the resolution of the Delta wave through to January 2023. This covered several significant immunological events including the Omicron BA.1 wave/peak third vaccine dose roll out (beige), third/fourth vaccine dose roll out (purple), Omicron BA.5 wave (red), Omicron BA.4/5 bivalent vaccine roll out (green) and Omicron BQ.1 wave (yellow). Cumulatively the pooled Privigen IVIG batches represent approximately 700,000 plasma donations. Time points are the date of plasma donation assigned to each IVIG batch and this is shown as months from the start of the COVID-19 pandemic. Average waste water SARSCoV-2 viral load (green line) in the U.S. population over that same period of time (courtesy of Biobot Analytics: https://biobot.io/). **B.** Live Virus Neutralization Titers (LVNT) of pooled IVIGs to ancestral Clade A.2.2 (black line; presented as endpoint titers normalized to WHO units/mL) overlaid with COVID-19 vaccine doses (red line) reported over that same period (https://ourworldindata.org/). **C.** LVNT (black line) versus anti-nucleocapsid IgG (red dashed line) in Privigen batches described in A. **D.** LVNT (black line) versus anti-Spike MFI levels (orange dashed line; measured using flow cytometry) in Privigen batches described in A. **E-F.** Correlation of LVNT (presented as WHO units/mL) to **E.** Anti-nucleocapsid IgG or **F.** Anti-Spike IgG. Each data point represents a Privigen batch. MFI = mean fluorescence intensity.

### Maturation of cross-reactive viral neutralization breadth throughout the COVID-19 pandemic overcomes initial immunological imprinting

Numerous studies have demonstrated that neutralization titers induced by COVID-19 vaccination and/or infection, significantly waned over time and as such led to re-infections with emerging circulating variants [10–13]. Concurrently, both ourselves and others have observed the decline in titers to be countered by increasing breadth and thus, an increase in the quality of the neutralization response [9]. To track the breadth of a population over several years, we determined LVNT for all major SARS-CoV-2 variants that appeared over the last three years using IVIGs from 2020 to the beginning of 2023. We focused on variants that sustained a threshold of greater than 50% of prevalence in the jurisdiction of the donor population (U.S. plasma donors) (Fig. 3A). Using ancestral Clade A.2.2 as the reference, LVNT of IVIGs to Delta, Omicron BA.1, BA.2, BA.5, BQ.1.1 and XBB.1.5 was then observed. Detectable titers were sustained across all other Omicron variants tested, albeit at levels lower than the earlier circulating pre-Omicron variants (Fig. 3A-B). Interestingly, a subsequent increase in titers to all variants was consistent with increased cross-reactivity over time. For calculation of breadth across isolates, we compared fold reductions in titers relative to the ancestral Clade A.2.2 (Fig. 3C). Throughout the pandemic, titers to the ancestral Clade A.2.2 have remained the highest among all variants over time, with the largest case wave of Omicron BA.1 failing to reverse this observation. The latter is consistent with a population that has sustained prior high vaccine uptake and/or experience of an earlier circulating variant infection and this is supported in prior observations of initial immunological imprinting in various cohort settings [14–19]. Whilst we observed results consistent with initial imprinting, this appears to be negated over time with increases in breadth across divergent variants. With the exception of Delta (with neutralization levels similar to Ancestral from early time points), the rate of increase in breadth over time, was similar across all variants (breadth linear slope: BA.1 = 0.03159; BA.2 = 0.03264; BA.5 = 0.02449; BQ.1.1 = 0.03202; XBB.1.5 = 0.04002. Mean slope of all variants 0.032 +/− 0.0055), which supports an absence of preferential targeting following any infection wave or vaccination period. The main difference between variants was the relative primary set points in viral titers of IVIG batches derived at the time of the resolution of the Delta infection wave (Fig. 3C). To estimate the trajectory of breadth, we applied linear regression to the breadth over time (where fold reduction of titers to ancestral Clade A.2.2 is 1). Here we started using IVIGs derived in early 2021, as they were the earliest samples to sustain LVNT across all variants. Using this approach, we estimated the time taken to reach equivalent neutralization breadth to ancestral for Omicron BA.1, BA.2, BA.5, BQ.1.1 and XBB.1.5 under the assumption that the slope would remain unchanged. With the starting timepoint at January 2023 (the last IVIG batch tested), the times ranged from approximately 0.33 years for BA.1 through to 2 years for XBB.1.5 (Fig. 3C). Given the rate of increase in breadth (slope of the linear regression in Fig. 3C) is similar across variants, we took the mean slope across all variants (0.032 +/− 0.0055) to establish a singular rate in the increase in IVIG breadth to all variants. This established the mean positive trajectory of antibody responses over approximately two years of the pandemic. Rather than focus on breadth of IVIGs over time, we analysed the increasing resistance to neutralization over time with the appearance of each variant. For this we plotted dominant variants from Delta through to EG.5.1 and their increasing resistance as calculated by the fold drop in neutralization compared to the ancestral variant Clade A.2.2 for a given IVIG batch. We then applied a linear regression and calculated the negative slope using this approach. In doing this our aim was to establish a rate of variant antibody evasion against all IVIG batches (an example for one IVIG batch from January 2023 is presented in Fig. 3D). In applying this analysis approach, we observed each IVIG batch over time to have a decreasing negative slope for variant evasion (Fig. 3E) (i.e. the more recent the IVIG batch the subsequent increase in viral resistance to neutralization over time was lower and as such the linear slope decreased over time; Fig. 3F). Under the assumption that the rate of antibody breadth increases and the rate of viral evasion remained constant, we estimate that in June 2024, the rate of breadth will equal and then surpass that of variant evasion (Fig. 3F).

**Figure 3.**
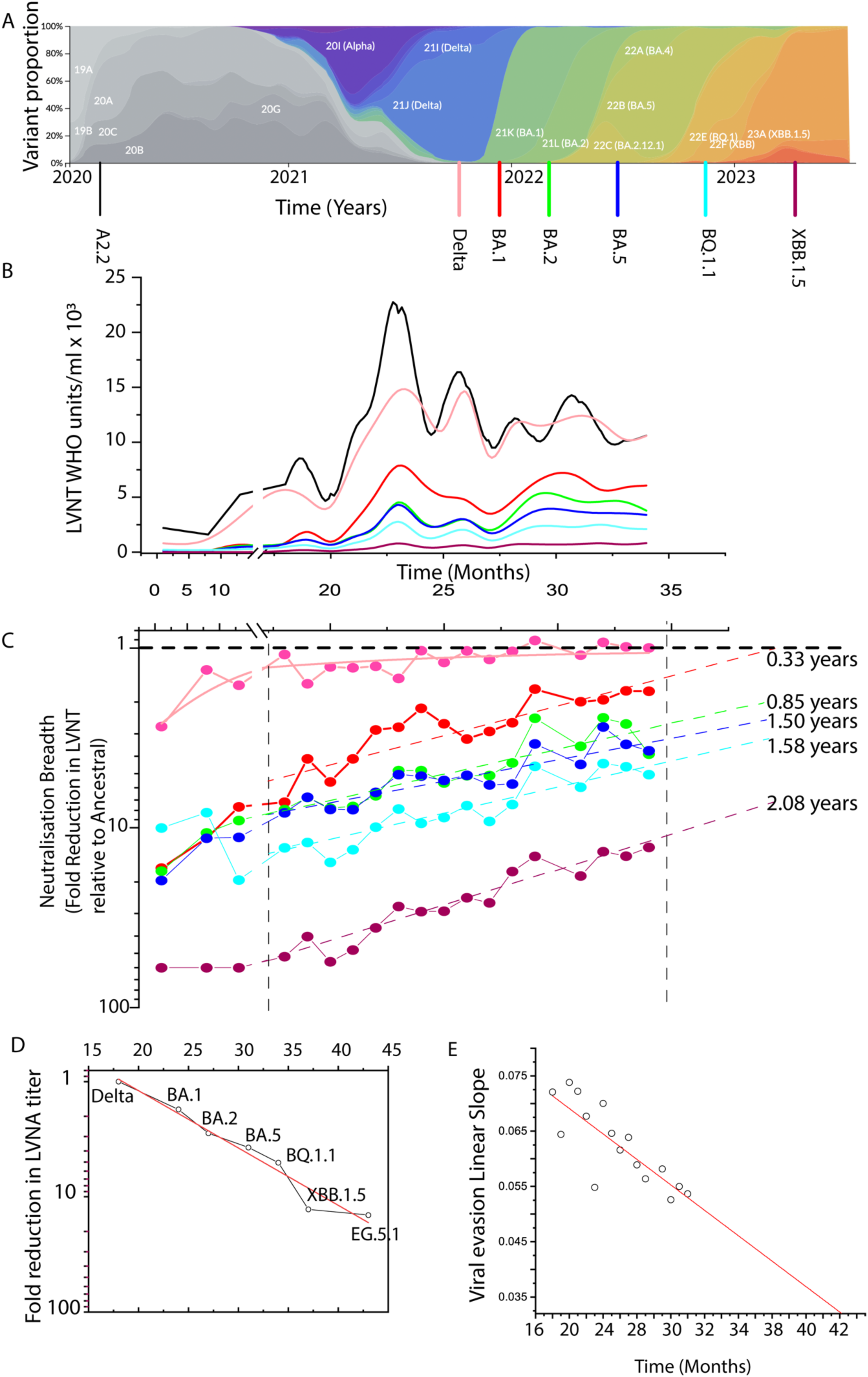
Increase in neutralization breadth over the first three years of the COVID-19 pandemic. **A**. Global frequency of SARS-CoV-2 variants are presented throughout the period of early 2020 – mid 2023 (nextstrain.org). Ancestral and major variants of concern that reached a threshold of greater than 50% prevalence in the U.S. population were selected for downstream testing. These isolates included an ancestral Clade A.2.2, Delta, Omicron BA.1, BA.2, BA.5, BQ.1.1 and XBB.1.5. **B.** Live Virus Neutralization Titers (LVNT) of pooled intravenous immunoglobulins (IVIGs) are presented as WHO Units/mL. The number of U.S.-based plasma donations collected across the time period outlined in A. represents approximately 720,000 donations. Each colored line represents LVNT to a SARS-CoV-2 variant outlined in A. **C.** Breadth over time to each SARS-COV-2 variant. Each data point represents the average LVNT for all pooled IVIGs batches collected within a single month. Data is presented as fold reduction of each variant (LVNT of Ancestral/LVNT of Variant) over time. The initial three points derived from IVIGs collected in 2020 did not reach titer to XBB.1.5 and for presentation fold reductions listed at 60-fold represent beyond the limit of detection for these data points. Calculated dashed lines represent the range of dates that are used to calculate breadth trajectories through linear fit curves for each SARSCoV-2 variant. The dashed lines for the breadth are then continued using the slope established from late 2021 through to early 2023 (vertical dashed black lines) to obtain approximate estimates of the time taken for maturation of the response to reach the equivalent of ancestral Clade A.2.2 (i.e. Fold reduction approaching 1). **D.** Representative example of the rate of decrease in LVNT of a given IVIG batch (January 2023) through the chronological appearance of each SARS-CoV-2 variant (black line) throughout the COVID-19 pandemic. A negative trajectory is observed with this batch through the loss of titer with the appearance of each variant over time (slope = –0.0498). **E.** Summary of the trajectory of each IVIG batch over time. With each IVIG batch (represented by a white dot), the slope of the variant evasion curve becomes lower as the breadth of each batch increases (the drop in titer for the appearance of each variant decreases). Assuming the evasion rate of the virus over time follows this trend and the rate of increase in IVIG breadth is also constant, then breadth gains in IVIG neutralization will equal evasion rates 18 months following January 2023 (June 2024).

### Emergence of JN.1 in late 2023 and dominance over XBB.1 sub-lineages is not associated with significantly decreased neutralization responses

In Australia in late 2023, XBB-derived sub-lineages dominated and were characterized by convergent polymorphisms accumulating at the type I antibody binding site. These included L452R/M/W (orange), L455F/S (green), F456L (pink) and F486P (blue) (Fig. 4A-B). Many XBB lineages acquiring the aforementioned convergent mutations were ranked high in global growth advantage, with the recombinant XCT gaining three type I antibody convergent polymorphisms (Spike positions 452, 455 and 456). At this time, there was low prevalence of a new divergent lineage BA.2.86, that was initially detected in Denmark and Israel [20]. Many BA.2.86 lineages were ranked highly in growth advantage over XBB sub-lineages, with a clear projected dominance for the BA.2.86 sub-lineage JN.1 defined by the L455S Spike polymorphism. As per our approach in early 2023, we ranked evasion IVIG batches consisting of the most recent donations at that time and tested viral evasion at the individual donor level in parallel. As we observe maturing breadth of antibody responses over time it was imperative that we utlised recently sourced donations. Unlike early 2023, the clear dominance of JN.1 in estimated growth rates and the convergence of XBB lineages with similar Spike polymorphisms (Fig. 4B-C) provided testing conditions that did not require significant triaging through large variant panel testing. Rather, using the most recently manufactured IVIGs (approximately June 2023) we tested a smaller SARS-CoV-2 variant panels, reflecting prior and current dominant variants circulating globally. This panel of variants included the ancestral Clade A.2.2, Delta, Omicron BA.1, BA.2, BA.5, BQ.1.1, XBB.1.5, EG.5.1, XCT and JN.1. In this panel, EG.5.1 induced the lowest titers with XCT and JN.1 exhibiting similarly low but not significantly different titers (Fig. 4D).

**Figure 4.**
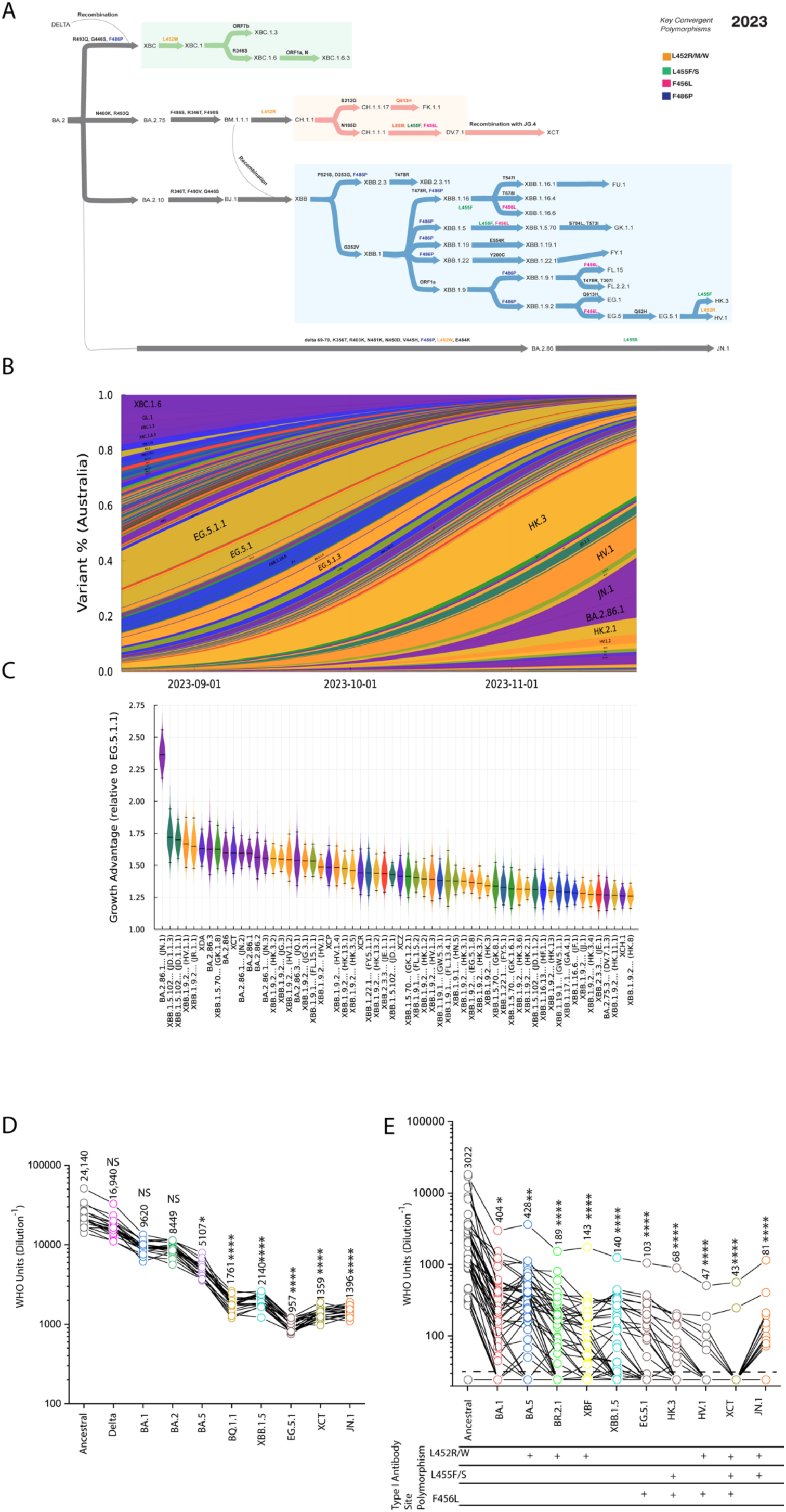
Neutralizing titers to emerging SARS-CoV-2 variants in late 2023. **A**. From left to right, Omicron lineages from the parental BA.2 to reflect the appearance of emerging variants in Australia by late 2023. There are three main groups from top to bottom: the upper grouping represents the Delta-BA.2 recombinant lineages XBC followed by the CH.1.1-derived lineages which now includes the recombinant XCT. The third group from the top is primarily XBB recombinant lineages with the emergence of HV.1 and HK.3 derived from EG.5.1. Convergent Spike polymorphisms across lineages are presented in blue (F486P), pink (F456L), green (L455F/S) and orange (L452R/M/W). The divergent BA.2.86 and its sub-lineage JN.1 is presented with key polymorphisms presented at the bottom for comparison. **B.** Surveillance summary of Australia in June 2023 based on genomic data via GISAID. Each vertical slice depicts the posterior mean variant frequency estimate from a hierarchical Bayesian multinomial logistic model of variant competition. Note the two dominant variant lineages are derived from EG.5.1 (HK.3 and HV.1; orange) with the latter appearance of BA.2.86 and JN.1 (purple) **C.** Results from a model of global SARS-CoV-2 lineage competition. Estimates of multiplicative growth advantage (per week) for lineages are provided, both relative to the parental BA.2, and to the recently dominant EG.5.1. D-E. Live virus neutralization titers were determined on **D.** Australian Lifeblood individual plasma donor samples collected primarily in the latter half of 2023 (n = 44) and **E.** 10 pooled intravenous immunoglobulins (IVIGs) manufactured from U.S. plasma donations from early to mid 2023. Inhibitory concentration 50 (IC50) values in **D-E** were calculated for each variant and compared to the ancestral Clade A.2.2. *** = P <0.001; **** = P<0.0001. Significance testing was performed using Freidman’s test with Dunn’s multiple comparison. Interpolated IC50 values were determined using the sigmoidal 4PL model of regression analysis in GraphPad Prism software version 9.1.2.

In parallel to testing of IVIGs representing population level immunity, we sought to test individual donors from previously curated cohorts. However, with many of these cohorts now concluding, a pragmatic source of recent donations linked to immunological histories came from the plasma donation program at our national blood bank, Lifeblood. Fortunately, the vaccination/infection profiles of these donors are representative of the immunological experiences of those within Australia and globally (Supplementary Table S1). We tested these individual donor samples against 11 SARS-CoV-2 variants selected based on their prior and current circulation in Australia. This panel included ancestral Clade A.2.2, Omicron BA.1, BA.5, BR.2.1, XBF, XBB.1.5, EG.5.1, HK.3, HV.1, XCT and JN.1. XCT displayed the lowest titers closely followed by HV.1, whereas HK3 and JN.1 had similar median titers to the other common circulating variants such as EG.5.1 and XBB.1.5 (Fig. 4E). This was in contrast to the higher LVNT sustained against ancestral and dominant Omicron lineages circulating in Australia in 2022 (BA.1, BA.5, BR.2.1 and XBF). These data show pooled IVIGs and individual donor plasma samples have significant similarities in their responses to SARS-CoV-2 variants. They also show that, unexpectedly, the neutralization responses alone could not explain the significant projected growth advantage of the JN.1 lineage compared to othe equally evasive XBB sub-lineages such as HK.3 and XCT.

### Divergence of JN.1 versus other sub-lineages is marked by preferential use of TMPRSS2 cleavage-resistant ACE2

Through the continued study of evolving entry pathways for emerging SARS-CoV-2 lineages, we have mapped a trajectory of ACE2 usage from the earliest ancestral Clade A through to contemporary Omicron lineages [7]. As SARS-CoV-2 also relies on Spike subunit 2 (S2) activation through TMPRSS2, we focused our studies on ACE2 use in the setting of ACE2 and TMPRSS2 co-expression. Through this approach, we have identified and engineered clonal cell lines with high TMPRSS2 activity that leads to cleavage modifications of ACE2 within its neck region. This ACE2 cleavage event phenotypically links early Clade A SARS-CoV-2 lineages with SARS-CoV-1, as both are augmented in entry when ACE2 is cleaved [2] (see Fig. 5A. for a visual summary of TMPRSS2 cleavage of ACE2 and Spike). In contrast, in Omicron sub-lineages we observe signficantly lower replication in the presence of cleaved ACE2 alongside TMPRSS2 [7]. To initially determine the preferential usage of cleavage modified/unmodified pools of ACE2, we titrated JN.1 using clonal lines expressing high TMPRSS2 activity [7] with cleavage sensitive ACE2 (WT ACE2) and cleavage resistant ACE2 (NC-ACE2) [7]. In the cleavage sensitive ACE2 setting, we observed attenuation of viral replication at levels that exceed all prior SARS-CoV-2 variants. In contrast, in the presence of cleavage resistant ACE2, we observed augmentation at levels that trended higher but were not significantly higher than prior lineages (2-fold) (Fig. 5C-G). Plotting the use of cleaved ACE2 over time with respresentative primary isolates we have obtained in our phenotypic surveillance activity, we observe a trajectory of decreasing infectivity (Fig. 5G). Of note, JN.1 is a clear outlier with the lowest level of viral infectivity in the latter setting (Fig. 5G). To date, using this analysis of the evolution of viral entry reveals a pathway for SARS-CoV-2 entry requirements that has consolidated to the use of ACE2 pools that are not cleaved by TMPRSS2. Further, we observed no evidence of reversion to viral entry observed in the earliest circulating SARS-CoV-2 isolates, where entry is augmented in the presence of cleaved ACE2. To validate this observation in primary cultures, we expanded both XBB.1.5 and JN.1 in parallel and challenged air-layer differentiated upper respiratory epithelial cells using equivalent viral particle number input. Three days post infection, we observed a significant increase in JN.1 viral loads compared to XBB.1.5 (5F) and thus consistent with a phenotypic shift in entry that can sustain a greater competitive advantage.

**Figure 5.**
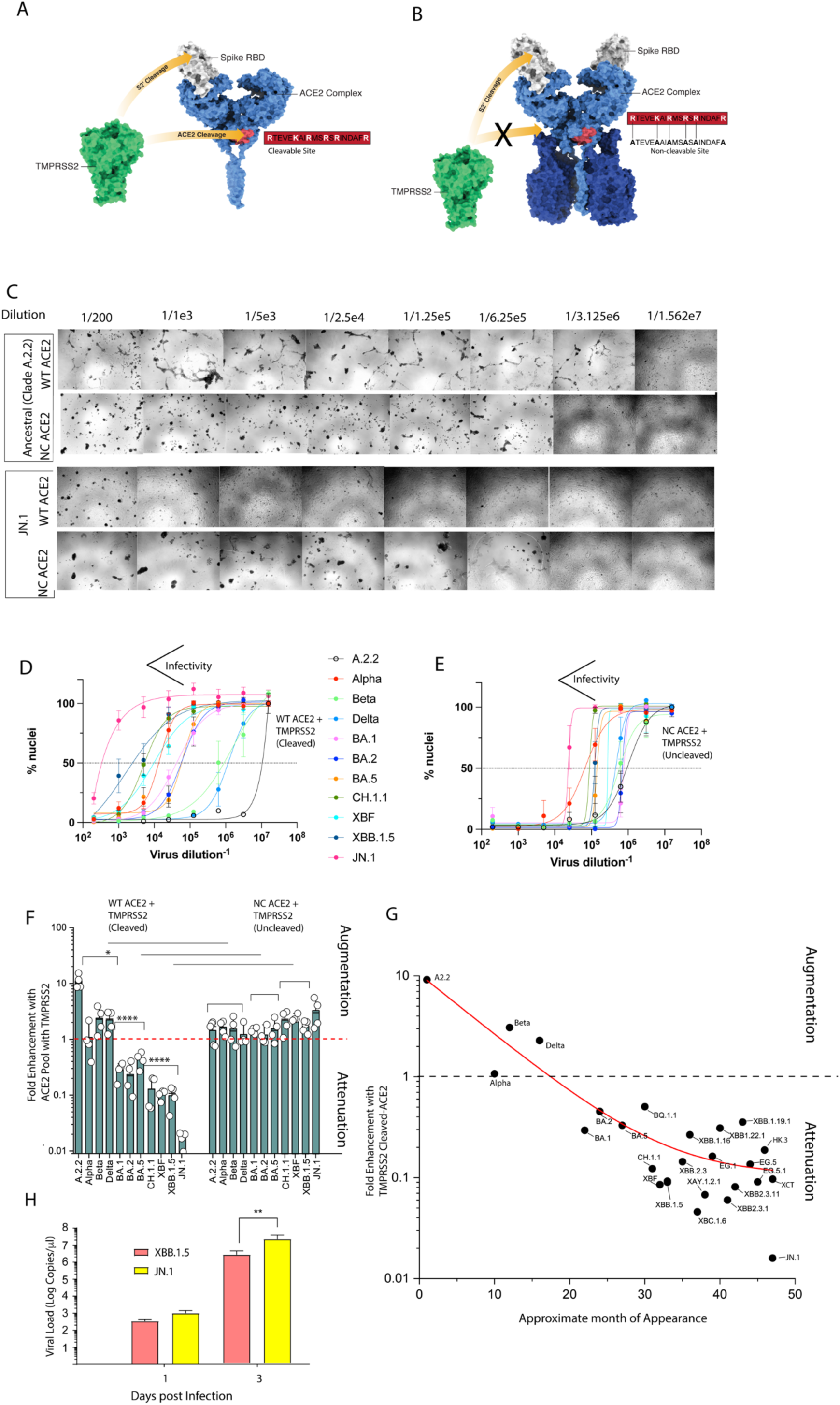
Preferential usage of TMPRSS2 cleavage resistant ACE2 over time and increased replication of JN.1 versus XBB.1.5 in primary upper respiratory epithelial cultures. **A**. The structure of the ACE2 dimer is presented (blue) based on the published structure [1], with the TMPRSS2 cleavage site (pink) and the cleavage sequence (boxed in red). TMPRSS2 (green) cleavage events within ACE2 and SARS-CoV-2 Spike glycoprotein (highlighted by arrows). **B.** The structure of the ACE2 dimer is presented as a dimer of heterodimers complexed with solute carrier SLC6A19 (dark blue) based on the published structure [1]. ACE2 complexes with either solute carriers SLC6A20 and SLC6A19 in and around the TMPRSS2 cleavage site, which is predicted to structurally occlude TMPRSS2 from accessing this site for cleavage. To control for lack of TMPRSS2 cleavage, an ACE2 cleavage resistant mutant (NC-ACE2) was generated, as previously described [2]. **C.** Both cleavage sensitive (WT ACE2) and cleavage resistant (NC-ACE2) were expressed alongside TMPRSS2 in the VeroE6 cell line. Both Ancestral Clade A.2.2 and JN.1 were titrated in both cell lines, with cytopathic effects presented after 2 days of culture. The top two rows of images demonstrate viral dilutions of Clade A.2.2 whilst the bottom two rows of images reflect JN.1 viral dilutions. **D. & E**. Representative titration curves using cells expressing **D.** wild type ACE2 and **E.** TMPRSS2 cleavage resistant ACE2 and TMPRSS2. For D. and E. viral replication is determined through dose dependent loss of live cell nuclei 3 days post infection. Standard deviations represent 8 technical replicates. **F.** Augmentation versus attenuation are presented for the major circulating variants throughout the pandemic (ancestral Clade A.2.2, Alpha, Beta, Delta, Omicron BA.1, BA.2, BA.5, XBB.1, CH.1.1, XBF, XBB.1.5, and JN.1). Titers are calulated here in the parental VeroE6-TMPRSS2 cell line alongside the WT ACE2 and NC-ACE2 cell lines shown in C. The red line signifies the point where the virus is replicating in the cell line at a level equivalent to its parent cell line VeroE6-TMPRSS2. * = P< 0.05 for decrease in attenuation in the NC-ACE2 versus WT ACE2 setting. **** = P<0.0005 for significant rescue in the NC-ACE2 versus WT ACE2 setting **G.** Augmentation and attenutation as outlined in **F.** presented with respect to time with representative lineages circulating in 2023. Decling use of cleavage sensitive ACE2 is presented (red line). **H.** Viral loads derived from primary ALI cultures. Clinical isolates XBB.1.5 and JN.1 were expanded and then equal RNA copies/ml was used to inoculate primary ALI cultures. Supernatants were harvested and RNA loads measured. Shown are the mean ± s.d. from *n* = 3 experiments ** = P<0.005.

## Discussion

As part of our continued SARS-CoV-2 surveillance, starting from early 2020 to late 2023, we combined cross-sectional observations and trajectories of longitudinal observations over the first 3 to 4 years of the COVID-19 pandemic. This encompassed the mapping of antibody responses in close to 1 million plasma donations and also resolution of primary isolate entry requirements. Using both approaches, we observed increased breadth in antibody coverage and an evolutionary trend of entry factor consolidation for variants towards a specific conformation of ACE2. Combined with cross-sectional studies of current variants, this establishes several pandemic “milestones” and conditions with contemporary variants that point to a positive breadth trajectory in antibody responses and a consolidation of viral entry pathway requirements that will importantly require greater resolution *in vivo* and further linkage to clinical observations. For recent cross-sectional work in 2023, rapid genotype to phenotype surveillance observed neutralization responses to JN.1 to be equivalent to many late circulating XBB lineages in 2023. This observation alone could not account for observed growth advantage of JN.1. In contrast, rapid *in vitro* growth assays revealed JN.1 to further consolidate its entry requirements to pools of uncleaved ACE2 co-expressed with the secondary entry factor TMPRSS2. This latter phenotype continues along the trajectory we have observed throughout the pandemic. Thus, we propose the ability of JN.1 to navigate antibody neutralization responses in addition to the consolidated and efficient usage of uncleaved ACE2 combined with TMPRSS2 are the primary means for its predicted and current growth advantage in the community.

We currently observe neutralization responses at the population level to have now plateaued following peak 3^rd^ dose vaccination doses and the overlapping large Omicron BA.1 wave. This is in face of decreasing vaccine booster uptake over time (https://ourworldindata.org/coronavirus) and community infections that were of similar magnitude to recent Omicron BA.5 and BQ.1 waves. The responses over time at the population level re-enforce observations from many cohort studies and this is observed at several levels. Firstly, continued high level responses to early circulating variants is consistent with immune imprinting with the initial ancestral Clade-based vaccine and/or convalescence from earlier circulating clades like Delta [14–19]. Secondly, resolution of cohort responses to breakthrough infections and/or newly developed vaccine formulations have yet to observe lineage-specific responses that result in targeting of a specific variant Spike glycoprotein [15, 21]. The most striking example of this is the dominant BA.1 wave, which alongside an overlapping vaccine roll out, drove the highest neutralization responses to the ancestral Clade and Delta. Nucleocapsid antibodies peaked around this time and this supported a large contribution of BA.1 infections to immune responses in the population. Similarly, the BA.5 infection wave also contributed to further peaks in Nucleocapsid antibodies, however this was not associated with the specific increases in LVNT breadth to BA.5 sub-lineages. Whilst we cannot rule out prior infections playing a role, the culminative observations aligning vaccine doses, case rates, wastewater viral loads and also Spike versus Nucleocapsid antibodies, collectively support vaccine doses as the dominant event that lead to increases in Spike antibodies, increases in LVNT and initial imprinting to the earlier ancestral clade. Fortunately the effects of early imprinting are now slowly reversing and this is demonstrated by increases in breadth to all variants at a rate similar over time. Given the equivalent gains in breadth across all variants, this is consistent with increasing cross-reactivity to all variants over time as observed at the individual donor level [21, 22]. The time taken for an approximate doubling in breadth is approximately one year (i.e. halving of fold reduction in LVNT relative to the ancestral strain), during which time new variants have emerged that counter these gains. This framed current cross-sectional surveillance in late 2023, where we observed many divergent variants such as HV.1, HK.3 and XCT accumulating several convergent Spike polymorphisms that could lower LVNT. Yet the dominance of XBB lineages ended upon the arrival of JN.1, which had growth estimates far in excess of all XBB-derived lineages in co-circulation in face of similar levels of neutralisation resistance [23–26].

At many stages of the pandemic, the acceleration in infection waves from a singular dominant variant was often associated with concurrent changes in entry requirements in addition to lowering of LVNT. This ranged from increases in ACE2 affinity [27] through to more efficient TMPRSS2 use like that observed with Delta and BA.5 [3, 28]. Whilst increasing gains in ACE2 affinity have been consistently observed in many studies [27, 29–35], the link to changing tropism of Omicron lineages from the lower to upper respiratory tract remains unclear. Through engineering of cell lines with differing ratios of ACE2 and TMPRSS2 [7], we have observed ACE2 modification through TMPRSS2 cleavage to significantly influence evolving viral entry over time and this provides a third entry requirement in need of careful consideration. Specifically, we observed the entry requirements of pre-Omicron lineages to significantly benefit from TMPRSS2 cleavage of ACE2 and this is also observed for SARS-CoV-1 [2]. In contrast, early Omicron lineages (BA.1 to BA.5) sustained attenuation when ACE2 was cleaved by TMPRSS2. Similarly, CH.1.1, XBC, XBF, XBB.1.5 and JN.1 were all characterized by decreasing infectivity after cleavage of ACE2, with the greatest drop observed in the latter JN.1 variant (Ancestral has 5 orders of magnitude higher titers than JN.1 with cleaved ACE2). In contrast, removal of the TMPRSS2 ACE2 cleavage site reverses attenuation and the fold increase is observed to be consolidated over time with the more recent Omicron lineages and especially JN.1. Compared to other recent studies, JN.1 and its parent BA.2.86 have generated a continuum of observations. Observations using primary clinical isolates across a number of cells lines has produced mixed results, with an unclear trajectory in tropism [24, 25, 36–39]. Further results of entry/fusion assays employing JN.1 and BA.2.86 pseudotypes produce results that diverge from those produced by JN.1 primary isolates [36, 38]. Yet the our observed entry phenotype is consistent with recent work published using primary isolates of JN.1 using similar cellular platforms to track viral replication [40]. Importantly, any surrogate assay for tropism needs to be linked to either clinical observations or where not possible *in vivo* observations in animal models. If the ACE2 cleavage model were to be predictive of lower respiratory tract tropism, early pre-Omicron isolates would observed efficient infection in the lung, whilst the progressive appearance of Omicrons would observe a trajectory of increasing attenuation. In recent studies comparing omicron BA.5, EG.5.1 and JN.1 in Syrian hamsters, they observe a rank order of attenuation BA.5<EG.5.1<JN.1 [41] and this is consistent with other independent studies the same animal model [37].

Physiologically monitoring viral replication and its sensitivity to ACE2 cleavage is consistent the physiological roles of ACE2 *in vivo* across various tissues. ACE2 cleavage is an essential regulatory element of the ACE2 release pathway in the renin angiotensin system, and it plays a well established role within the lower respiratory tract in response to acute injury [42]. Therefore the sensitivity of ACE2 to cleavage in that tissue is well known and there is now accumulating evidence to support these cleavage events may now be protective through reducing of viral replication in Omicron lineages within this tissue. Whilst the above is consistent with consolidated tropism away from the lower respiratory tract, efficient replication in other tissues would need to take place to be consistent with continued spread and dominance of linaeages like JN.1.

In other tissues, ACE2 acts as a chaperone for solute carrier (SLC) proteins such as SLCA619 and SLCA620. It forms a dimer of heterodimers in which the solute carrier is complexed with ACE2 (starting from ACE2 residue 621) at residues that may enable exclusion from the TMPRSS2 cleavage site (ACE2 residues starting from 697) [43, 44]. High levels of ACE2 and SLCA619 expression in enterocytes of the small intestine would support a TMPRSS2 resistant pool of ACE2 and the potential site of evolution for newly emerging variants like JN.1. Whilst replication kinetics in primary enterocytes have yet to be observed for JN.1, recent cohort studies of viral faecal shedding support continued and increased replication at this tissue site [45]. The latter does not readily explain the enhanced growth advantage within the upper respiratory tract. Whilst ACE2-SLCA619 chaperone activity is well established in enterocytes, ACE2-SLCA620 chaperone activity is also associated with the respiratory tract and genetic association studies have revealed single nucleotide polymorphisms in the SLCA620 gene that are primarily associated with the relative risk of infection [46, 47]. The latter is consistent with our observations in primary air differentiated bronchial epithelial cells, where we do see a shift in entry requirements in cells line consistent with greater replicative capacity when comparing XBB.1.5 and JN.1. Given the persistence of SARS-CoV-2 lineages and recent consolidation of JN.1 towards use of cleavage resistant pools of ACE2, we presently hypothesize that there is a transmission advantage in targeting ACE2 away from its role in the renin angiotensin system but rather in its chaperone function alongside solute carriers. Future *in vivo* studies will be paramount in understanding; (1) how the cleavage events of entry receptors may influence the fitness of emerging variants and, (2) whether they contribute to a change in tropism with a specific disease profile in the clinic.

In summary, continued cross-sectional and longitudinal surveillance of neutralization responses against dominant SARS-CoV-2 variants will be important in tracking the pandemic during this phase of high case waves alongside declining vaccine uptake within the majority of the population. Whilst break through infections have been observed to sustain good neutralization responses, the impact of continued infection waves within the community and the clinic will need to be monitored carefully. Alongside this clinical monitoring, tracking the entry requirements of past and emerging variants will remain vital including the ability of the virus to influence disease severity within the present immunological setting. Knowledge of the latter will give greater resolution with respect to viral tropism and the impact of acute versus chronic infection on various tissue types. Further study may reveal a trajectory towards our current experiences with related seasonal human coronaviruses that have been in circulation for decades [48]. Yet we need to remain vigilent and pragmatic surveillance approaches can enable rapid phenotypic observations of variants circulating in the community to provide feedback for appropriate recalibration of our public health and vaccine responses when need be. Furthermore, the longitudinal time courses across large populations observed within this work will provide a dataset that can be used in future pandemic responses in understanding the trajectory of a population and virus over the emergency phase of a pandemic.

## Contributors

Assay was developed and performed by A. Aggarwal, A. Akerman, C. Fichter and S. G. Turville. Viruses were isolated and propagated by A. Aggarwal, A. Akerman, and S. G. Turville. Experiments were performed by A. Aggarwal, A. Akerman, C. Fichter, C. Esneau, J. Lopez, V. Milogiannakis, J. Caguicla and S. G. Turville. Additional research support including whole genome sequencing, serum samples and pooled IgG batches were provided by Z. Naing, M. Yeang, A. Condylios, W. Rawlinson, S. Amatayakul-Chantler, N. Roth, S. Manni, T. Hauser, V. Sintchenko, J.Kok, D. Dwyer, E. Knight, D. Irving, I. Gosbell, V. Hoad, D. Darley, G.Mathews, Promsri S, Stark D. Linkage of phenotypic surveillance was facilitated by the NSW ministry of health through K. Marris and S. Ellis. Summary of Australian variant analysis and global competition analysis was performed by B. Murrell and K. Sato. Analysis of experiments was performed by A. Aggarwal, A. Akerman, C. Fichter, J. Wood, M Silva, T. Ison and S. G. Turville. Data was verified by A. Aggarwal, A. Akerman, C. Fichter and S. G. Turville. The manuscript was drafted by A. Aggarwal, A. Akerman, C.Fichter and S. G. Turville. The study was supervised by A. Aggarwal and S. G. Turville. All authors read and approved the final version of the manuscript.

## Data Sharing Statement

Source data for generating the figures are available in the online version of the manuscript. Any other data are available on request.

## Data Availability

All data produced in the present study are available upon reasonable request to the authors

